# Community engagement to increase vaccine uptake: Quasi-experimental evidence from Islamabad and Rawalpindi, Pakistan

**DOI:** 10.1101/2022.09.04.22279583

**Authors:** Mujahid Abdullah, Taimoor Ahmad, Twangar Kazmi, Faisal Sultan, Sabeen Afzal, Rana Muhammad Safdar, Adnan Ahmad Khan

**Author notes:** Corresponding author: (AAK).

## Abstract

Developing countries have been facing difficulties in reaching out to low-income and underserved communities for COVID-19 vaccination coverage. The rapidity of vaccine development caused a mistrust among certain subgroups of the population, and hence innovative approaches were taken to reach out to such populations. Using a sample of 1760 respondents in five low-income, informal localities of Islamabad and Rawalpindi, Pakistan, we evaluated a set of interventions involving community engagement by addressing demand and access barriers. We used multi-level mixed effects models to estimate average treatment effects across treatment areas. We found that our interventions increased COVID-19 vaccine willingness in two treatment areas that are furthest from city centers by 7.6% and 6.6% respectively, while vaccine uptake increased in one of the treatment areas by 17.1%, compared to the control area. Our results suggest that personalized information campaigns such as community mobilization help to increase COVID-19 vaccine willingness. Increasing uptake however, requires improving access to the vaccination services. Both information and access may be different for various communities and therefore a “one-size-fits-all” approach may need to be better localized. Such underserved and marginalized communities are better served if vaccination efforts are contextualized.

## Introduction

The SARS-CoV-2 (COVID-19) outbreak has been one of the largest infectious disease challenges in the past century with 559 million cases and 6.36 million deaths [1]. In the initial phase, the only means to curb transmission were measures that limited contact between individuals, such as lockdowns, closures of schools, work and public places. These, in turn resulted in tremendous social costs and loss of wellbeing of individuals and societies [2]. However, the availability of effective vaccines from the first half of 2021 changed how countries and societies approached the contagion and how effective they were in doing so.

The rapidity of the vaccine development process has been unprecedented, as has been the intended scope of its coverage. Until now it took 5-10 years to develop and navigate most vaccines through regulatory approvals, which for most parts were administered to sub-sets of population such as children, women giving birth etc. COVID-19 vaccines went from the first identification of the virus to a public rollout of vaccines in under one year and were aimed at nearly all of the world’s population.

This swiftness raised issues of mistrust among potential recipients, who questioned both the efficacy and safety of the vaccine. This in turn led to some reluctance and affected the rollout of the vaccine. Several means were attempted to promote vaccination widely, including mandates (e.g., for healthcare workers, other government personnel or certain patients) [3,4], tying vaccination to access to public transport or to enter stadiums, or by giving incentives to vaccinate (e.g., discounts on certain purchases). As anticipated, much of the initial vaccinations were in cities, and among well-documented and vaccine seeking populations. However, the large-scale rollout and the intent to cover the entire population required addressing several complex situations.

For one, globally, about 1 billion people reside in densely populated low-income informal settlements (urban slums) [5] where access and availability to public health facilities is limited [6–8]. These barriers are further accentuated by a mutual lack of trust between these residents and public authorities that are supposed to serve them, but often do not have a full sense of their numbers, nor have systems in place to reach and meet their needs, and often consider them illegal occupants of government lands [9,10]. This in turn creates social exclusions and aggravates intra-societal iniquity where the most marginalized individuals are also suspicious of the government and its initiatives to reach them with life-saving services [11–15].

Pakistan started a multi-staged rollout of COVID-19 vaccination in March 2021 that initially prioritized the oldest population, frontline workers, and those with certain risk factors, and then progressively included younger citizens, till vaccinations were opened to everyone aged 18 years or older from July 2021. When the original, voluntary uptake of vaccines slowed down, several strategies were attempted, including reaching out to poor urban communities [16]. The present study explores the effectiveness of interventions aimed at addressing demand and access barriers in such communities.

## Theoretical framework for the study

Several socio-demographic factors, communication about COVID-19 and vaccines, perceptions regarding COVID-19 vaccination, and prior experience of COVID-19 infection affect vaccine acceptance. Being male, older in age, highly educated, and employed are associated with higher acceptance; as is the perception of COVID-19 risk towards oneself and a personal or family’s history of COVID-19 infections [11,17– 26]. In addition, information and communication about COVID-19 vaccination act as signals to influence individual behavior [24,25].

These factors were included in the context of the Theory of Planned Behavior (TPB) to develop a theoretical framework of vaccine willingness and uptake. The theory suggests that people’s behavioral intentions are motivated by their attitude, subjective norms, and perceived behavioral control [27]. These behavioral intentions in turn can directly affect an individual’s health behavior [18,27,28], which we hypothesized in our study as willingness translating to increased vaccine uptake among the targeted population.

Through our community engagement interventions, we aimed to change behavioral intentions (i.e., COVID-19 vaccine willingness) of residents in the treatment areas. Behavior change communication was carried out in context of social mobilization to engage the communities [29–31]. Interventions aimed at mobilizing communities for vaccination can help strengthen weak links in the causal chain, as this can enable to take into account the local characteristics and implement the interventions in a more effective way [32]. When information is spread through local prominent members (community and religious leaders) of the community, people become more willing to accept it and in turn, implement it.

Engaging communities thus aid to disseminate information in the local language and channels which can have a greater outreach [30]. As found in an earlier study of rural Bangladesh [33], collaboration with NGOs to increase immunization rates also results in better service delivery and increases vaccination acceptance as people exhibit more trust in local NGOs. Mobile vaccination camps (MVCs) can help increase access to vaccinations in such underserved communities. The current study explored the roles of community mobilization and vaccination camps in marginalized, low-trust communities to promote awareness and uptake of COVID-19 vaccination and the effects of such interventions in sub-populations of these communities.

## Methodology

### Study overview, location and sampling

The study used a cross-sectional research design. Residents of five urban poor communities from Rawalpindi-Islamabad twin cities were included. A baseline survey of 1760 respondents with equal representations of males and females was conducted from June 16 to 26, 2021, followed by an intervention (explained below). An endline survey was conducted from August 24 to September 03, 2021, with the same sampling technique, but not the same respondents. The response rate for baseline was 98% while it was 96% for the endline.

The study was limited to COVID-19 vaccine-eligible respondents of 18 years of age that were residents of selected communities. The final survey instrument comprised of 38 questions divided into multiple sections. Only a few questions were open-ended and the survey was administered in Urdu (local) language for accurate responses. Data collection was carried out in field on electronic tablets using SurveyCTO.

Five densely populated, low-income, and underserved areas were selected in consultation with the Ministry of National Health Services, Regulations and Coordination (MoNHSRC) for their low participation in the ongoing vaccination efforts. From Islamabad, we included I-10 (a middle-class locality), G-7 (Low-income but formal locality), F-7 (France Colony) (informal settlement), and Bhara Kahu (low-to middle-income, completely informal and recent settlement), while Dhok Hassu (low-income, long stand informal locality) was included from Rawalpindi. Each community is located sufficiently away from each other to make any cross over contamination of the intervention (information and eased access to local camps through social mobilization) unlikely.

Population and average household sizes were based on the census 2017 by Pakistan Bureau of Statistics and on-site visits (Table 1).

**Table 1.** Location characteristics of study areas.

| Locality | Actual Population | Actual Households | Average Household Size |
| --- | --- | --- | --- |
| I-10 | 44,580 | 7,984 | 5.6 |
| G-7 (Low-income quarters) | 29,609 | 4,707 | 6.3 |
| F-7 (France Colony) | 9,113 | 1429 | 6.4 |
| Bhara Kahu | 125,048 | 21,123 | 5.9 |
| Dhok Hassu | 201,212 | 30,032 | 6.7 |

The sample size was calculated using MICS methodology with a 95% confidence interval [34]. We assumed a 50% acceptance rate for vaccination uptake, a design effect of 1.5, a relative margin of error of 0.12 and a 95% response rate. The sample size included 480 respondents from each of the larger communities (population greater than 30,000), while 160 each from smaller communities.

A two-stage clustered sampling design was applied using GIS mapping, with randomization being done first by selecting random sample of clusters in each locality of the sampling frame and then at selecting households from each cluster. Pins identifying clusters were dropped at random points on map. Out of those, we randomly selected a total of 110 clusters, 30 each in Dhok Hassu, Bhara Kahu and I-10, and 10 each in F-7 (France Colony) and G-7 (Low-income quarters) for each round of surveys.

Working in pairs, enumerators reached the pins and started with the nearest household to the left within the cluster. The pair then surveyed fifteen more households in that cluster using the left-hand rule, skipping one household after every successful interview. Each pair surveyed 8 male and 8 female respondents in every cluster, for a total of 16 interviews per cluster. Males and females were surveyed from different households, with no upper age limit restriction.

## Interventions

### Awareness campaign via local mobilizers

The primary intervention focused on building awareness among residents of treatment areas through social mobilization techniques geared towards improving vaccine willingness and uptake. The campaign targeted public places such as shops, markets, mosques and churches. Within the communities, active members of the community including religious and political leaders were engaged to spread awareness on COVID-19. Printed information pamphlets (S1 Fig) were also distributed in Urdu language to explain the process of registering and getting vaccinated, while also debunking common myths surrounding vaccines, and identifying COVID-19 vaccination camps (CVCs) nearby.

### Mobile vaccination camps (MVCs)

Since there were no CVCs in the vicinity of the selected communities, MVCs were arranged to provide access to COVID-19 vaccination. These camps were organized in treatment areas in collaboration with local community-based organizations, NGOs and local leaders through a team of volunteers (usually local activists) to facilitate the community vaccination process encompassing assembling, counselling and registering community members for the vaccination. The venue of the vaccination site was chosen by the community as a locally well-known and accessible location, such as a school or other landmarks. Local volunteers also advertised for these in advance and on the day of the visit, they helped improve access of community members to the camps and facilitated them while they were at the camp.

## Empirical measurement strategy

We used intent-to-treat (ITT) analysis to measure average treatment effects (ATEs), assuming that households remained in the same treatment groups to which they were originally assigned, whether they received the treatments or not. We estimated ATEs on two primary outcomes: willingness to vaccinate and vaccine uptake. Given both the dependent variables were binary, we estimated non-linear ITT parameters using multi-level mixed effects logistic regressions [35] through difference-in-differences method. Here, level 2 indicates clusters and level 1 indicates households within those clusters. As suggested by Bruhn and McKenzie [36], we did not report statistical differences between groups at baseline covariates.

The control variables in our analyses were taken based on the theoretical framework explained above as well as their predictive powers to explain the outcome variables [36], which were calculated as having strong correlations with the outcome variables. Controlling for these variables that could be imbalanced at the baseline also controlled for imbalance in the unobservable characteristics [36], and therefore the difference-in-differences analysis was applicable. We ran all our analyses on statistical software STATA 17.

The difference-in-differences model in regression form is then specified as follows:

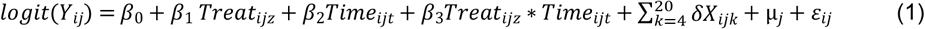

In equation 1, *i* refers to each household, *j* refers to each cluster, z represents each treatment group, *t* represents pre-and post-time periods and *Y*_*ij*_ is the relevant outcome. Since we estimated two models, *Y*_*vu*_ was vaccine uptake and *Y*_*wv*_ was willingness to vaccinate. We also accounted for several control variables 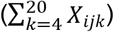 in our models to explain variation in our outcome variables (S1 Table). The fixed part of the model consists of 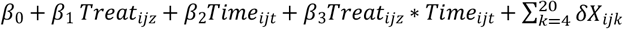 represents the random part of the model, and ε _ij_ is the household-level specific error term.

Vaccine uptake (*Y*_*vu*_) is given a value of 1 if the respondent has received at least one dose of COVID-19 vaccination and 0 otherwise, and willingness to vaccinate (*Y*_*wv*_) is assigned a value of 1 if the respondent is willing to get vaccinated if a free of cost government-administered COVID-19 vaccine is provided, and 0 otherwise. *Treat*_*ijz*_ indicates the localities which we took in treatment and control areas. We define three treatment areas (T1: G-7/F-7, T2: Bhara Kahu and T3: Dhok Hassu) and one control area (C: I-10). *Time*_*ijt*_ is a binary variable indicating a value of 1 for post-intervention and 0 for pre-intervention time periods.

The coefficient (*β*_*3*_) on *Treat*_*ijz*_ * *Time*_*ijt*_ captures the effect of interventions on the treated areas as compared to the control area. Since our model is non-linear in nature, ATEs are calculated by cross derivatives with respect to time and treat variables [37]:

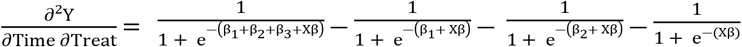

The R-square for our models were calculated using a community-distributed STATA program written by Dr Wolfgang Langer [38].

## Results

At the baseline, the respondents had a median age of 35 (range: 18-86) years. They were predominantly of Punjabi ethnicity, with the exception of Dhok Hassu, where 42% respondents were Pashtun (Table 2). The control area (I-10) had more respondents that were Urdu speakers (10%), were better off than any of the treatment areas, and more educated – fewest uneducated (9%) and the most university degree holders (47%). Unemployment rates ranged from 61% in I-10 to 48% in Dhok Hassu.

**Table 2.** Descriptive baseline characteristics in percentages across groups.

| Variables | Categories | Control Group |  | Treatment Groups |  |  |  |  |  |
| --- | --- | --- | --- | --- | --- | --- | --- | --- | --- |
|  |  | I-10 |  | G-7/F-7 |  | Bhara Kahu |  | Dhok Hassu |  |
|  |  | Baselin<br>e | Endlin<br>e | Baselin<br>e | Endlin<br>e | Baselin<br>e | Endlin<br>e | Baselin<br>e | Endlin<br>e |
| Age Group | 18-29 | 32.2 | 28.3 | 31.7 | 36.0 | 29.2 | 26.6 | 30.8 | 29.2 |
|  | 30-39 | 24.5 | 24.0 | 27.3 | 21.9 | 31.9 | 33.2 | 31.0 | 28.0 |
|  | 40-49 | 19.0 | 18.7 | 19.1 | 19.3 | 22.9 | 22.7 | 22.8 | 20.9 |
|  | 50-59 | 11.2 | 14.8 | 13.2 | 12.8 | 9.7 | 12.6 | 9.4 | 12.9 |
|  | 60+ | 12.6 | 14.3 | 8.8 | 10.0 | 6.3 | 5.0 | 6.1 | 9.1 |
| Education level | None | 8.8 | 6.3 | 19.5 | 24.6 | 15.8 | 15.9 | 34.5 | 34.6 |
|  | Up to 12 years | 44.0 | 42.3 | 57.9 | 60.1 | 63.0 | 64.5 | 58.5 | 58.1 |
|  | University degree | 47.3 | 51.4 | 22.6 | 15.3 | 21.2 | 19.6 | 7.1 | 7.2 |
| Ethnicity | Punjabi | 58.0 | 59.1 | 84.0 | 90.7 | 50.6 | 63.9 | 45.1 | 48.7 |
|  | Pashtun | 16.74 | 15.2 | 5.6 | 4.7 | 16.2 | 13.6 | 42.4 | 39.0 |
|  | Urdu Speaking | 10.3 | 16.1 | 2.2 | 0.6 | 3.4 | 5.0 | 0.4 | 0.62 |
|  | Hindko | 3.6 | 2.7 | 1.3 | 0 | 4.4 | 3.1 | 6.9 | 7.2 |
|  | Others | 11.5 | 7.0 | 6.9 | 4.1 | 25.4 | 14.4 | 5.2 | 4.5 |
| Employment | Self Employed | 15.7 | 11.9 | 6.9 | 10.3 | 19.3 | 21.3 | 29.4 | 25.0 |
|  | Employed | 23.1 | 22.8 | 39.8 | 38.6 | 26.9 | 22.1 | 22.4 | 21.4 |
|  | Unemployed | 61.2 | 65.3 | 53.3 | 51.1 | 53.8 | 56.6 | 48.2 | 53.6 |
| Self-infection of COVID-19 | Yes | 13.5 | 13.7 | 7.3 | 4.4 | 4.3 | 3.5 | 1.5 | 2.7 |
| Family infection of COVID-19 | Yes | 16.9 | 19.7 | 11.6 | 6.0 | 7.0 | 6.4 | 2.3 | 3.3 |
| Sought treatment for last illness | Yes | 79.3 | 72.2 | 79.6 | 70.0 | 80.6 | 74.8 | 83.3 | 81.4 |
| Family vaccination | Yes | 57.4 | 85.5 | 66.9 | 89.3 | 36.4 | 69.0 | 18.9 | 57.0 |
| Distance from CVC | Up to 2 kms | 5.0 | 23.3 | 45.3 | 20.6 | 43.9 | 49.6 | 8.2 | 55.5 |
|  | More than 2 kms | 49.4 | 40.3 | 42.8 | 67.6 | 29.4 | 28.1 | 37.1 | 22.3 |
|  | Don't Know | 45.6 | 36.4 | 12.0 | 11.8 | 26.7 | 22.3 | 54.6 | 22.3 |
| Risk perception of COVID-19 | Worried | 65.8 | 68.5 | 63.6 | 61.7 | 59.0 | 77.9 | 71.4 | 74.1 |
|  | Uncertain | 17.2 | 10.9 | 4.8 | 6.2 | 14.8 | 9.3 | 10.7 | 9.1 |
|  | Unworried | 17.0 | 20.6 | 31.7 | 32.1 | 26.2 | 12.8 | 17.9 | 16.8 |
| Sources of information: |  |  |  |  |  |  |  |  |  |
| Television | Yes | 26.6 | 35.0 | 33.2 | 24.6 | 25.4 | 14.2 | 23.4 | 28.3 |
| Government Call/SMS | Yes | 26.8 | 17.8 | 16.3 | 19.9 | 37.6 | 41.0 | 27.6 | 32.4 |
| Family/Friends | Yes | 46.2 | 24.2 | 49.2 | 53.0 | 46.0 | 65.2 | 44.1 | 63.1 |
| Medical professionals | Yes | 8.6 | 8.4 | 9.1 | 18.1 | 6.1 | 11.6 | 4.2 | 15.9 |
| Religious leaders | Yes | 0 | 0 | 0.3 | 7.5 | 1.5 | 2.7 | 0.4 | 6.2 |
| Any NGO/CBO working in area | Yes | 88.7 | 66.6 | 56.1 | 48.3 | 74.4 | 71.6 | 74.1 | 73.4 |

Few respondents (1% to 17%) reported any prior COVID-19 infection for themselves or among their families from any location. The highest rates were reported from the control area (I-10). However, 59-71% of all respondents reported being worried about COVID-19. The history of at least one member of the family having received the COVID-19 vaccine was the highest in G7/F7 (67%), followed by I-10 (57%), and the lowest in Dhok Hassu (19%). Distance from a CVC was the least for residents of G7/F7 and Bhara Kahu and the most for I-10 residents. Respondents from all areas reported similar proportions of sources of COVID-19 vaccination information and similar rates of treatment seeking during a prior illness. 89% of respondents from I-10 were aware of NGOs and CBOs working in the area, compared with 56% in G-7/F-7 and 74% in Bhara Kahu and Dhok Hassu (Table 2).

### Willingness to vaccinate and COVID-19 vaccine uptake

Willingness to receive vaccines increased substantially from baseline (67%) to endline (80%), more for men than women. The control area (I-10) had the highest willingness for both men and women across both time periods, with an exception that G7/F7 had the highest willingness for men at the endline (94%).

Correspondingly, refusal to receive vaccine dropped sharply in the endline. For men, the highest dip in refusals occurred in Dhok Hassu (18%), while Bhara Kahu had the highest decrease for women (20%). Registrations and vaccinations mirror willingness. Vaccine uptake increased from 22% to 47%, with men receiving more vaccination than women (Table 3).

**Table 3.** Willingness to vaccinate and vaccine uptake segregated by gender and location.

|  |  | Willingness to vaccinate |  |  |  |  |  | Vaccine uptake |  |  |  |  |  |
| --- | --- | --- | --- | --- | --- | --- | --- | --- | --- | --- | --- | --- | --- |
|  |  | Willing |  | Uncertain |  | Unwilling |  | Unvaccinated |  | Only registered |  | At least partially vaccinated |  |
|  |  | Baseline | Endline | Baseline | Endline | Baseline | Endline | Baseline | Endline | Baseline | Endline | Baseline | Endline |
| **TOTAL** |  | 67% | 80% | 12% | 9% | 21% | 10% | 65% | 39% | 13% | 14% | 22% | 47% |
| **Male** | C: I-10 | 82% | 88% | 9% | 2% | 9% | 10% | 59% | 20% | 12% | 31% | 29% | 49% |
|  | T1: G-7/F-7 | 81% | 94% | 2% | 4% | 17% | 2% | 43% | 16% | 16% | 6% | 40% | 78% |
|  | T2: Bhara Kahu | 64% | 77% | 25% | 18% | 12% | 5% | 64% | 35% | 12% | 14% | 24% | 51% |
|  | T3: Dhok Hassu | 64% | 85% | 13% | 10% | 23% | 5% | 76% | 38% | 14% | 22% | 10% | 40% |
|  | **Total** | 72% | 85% | 13% | 9% | 15% | 6% | 62% | 28% | 13% | 19% | 24% | 52% |
| **Female** | C: I-10 | 73% | 83% | 9% | 9% | 18% | 9% | 49% | 32% | 18% | 11% | 33% | 57% |
|  | T1: G-7/F-7 | 71% | 72% | 8% | 8% | 21% | 20% | 58% | 43% | 13% | 6% | 29% | 51% |
|  | T2: Bhara Kahu | 51% | 80% | 13% | 4% | 36% | 16% | 79% | 60% | 8% | 7% | 12% | 32% |
|  | T3: Dhok Hassu | 55% | 67% | 17% | 18% | 28% | 15% | 83% | 59% | 11% | 12% | 7% | 28% |
|  | **Total** | 62% | 76% | 12% | 10% | 26% | 14% | 68% | 49% | 12% | 9% | 20% | 41% |

### Intent-To-Treat (ITT) and Average Treatment Effects (ATEs)

Average marginal treatment effects from multi-level mixed effects logistic regressions show that, compared to the control area (I-10), willingness to receive vaccination increased by 7.6% in Bhara Kahu (Coef: 0.0764, CI: 0.0121, 0.1406) and 6.6% in Dhok Hassu (Coef: 0.0661, CI: 0.00498, 0.1272) respectively. However, the change in G7/F7 was not significant.

Whereas, this willingness did not translate into an increase in vaccination rates in either of the areas. Vaccine uptake increased by 17.1% (Coef: 0.1709, CI: 0.0417, 0.3) in G-7/F-7 only (Table 4). Our adjusted models with all controls showing odds ratios are provided in S2 Table.

**Table 4.** Pairwise comparisons of average marginal treatment effects.

| Comparison | (1)<br>Willingness to vaccinate |  | (2)<br>Vaccine Uptake |  |
| --- | --- | --- | --- | --- |
|  | Unadjusted | Adjusted | Unadjusted | Adjusted |
| **Difference-in-differences** |  |  |  |  |
| T1: G-7/F-7 vs C: I-10 | 0.0312<br>(-0.0578, 0.1203) | 0.0611<br>(-0.0197, 0.1419) | 0.077<br>(-0.0354, 0.1894) | 0.1709**<br>(0.0417, 0.3) |
| T2: Bhara Kahu vs C: I-10 | 0.1189***<br>(0.0424, 0.1954) | 0.0764**<br>(0.0121, 0.1406) | 0.0078<br>(-0.0994, 0.115) | 0.0418<br>(-0.0817, 0.1652) |
| T3: Dhok Hassu vs C: I-10 | 0.1314***<br>(0.0566, 0.2063) | 0.0661**<br>(0.00498, 0.1272) | 0.0385<br>(-0.066, 0.143) | 0.0398<br>(-0.0767, 0.1563) |
| Observations | 3107 | 2,904 | 3448 | 3216 |
| Number of clusters | 220 | 220 | 220 | 220 |
| Intra-class correlation | 0.044 | 0.046 | 0.042 | 0.048 |
| McKelvey & Zavoina R2 (FE and RE) | 0.083 | 0.395 | 0.198 | 0.533 |

Robust standard errors were used, CI in parentheses
\*\*\* p<0.01, \*\* p<0.05, \* p<0.1

The Intra class correlation (ICC) is the correlation among observations within the same cluster. In our models, ICC indicates that only around 4.2-4.8% of the total variance in willingness to vaccinate and uptake is explained by between-cluster differences (i.e., due to clustering). The Mckelvey & Zavoina Pseudo R-squares of adjusted models show that 40% and 53% of the variations in willingness and uptake respectively are captured by the independent variables. Both models show good fits to predict the relevant outcomes.

### Determinants of willingness and vaccine uptake

Willingness to vaccinate was twice as likely in the control area at baseline but this effect disappeared at the endline. On the other hand, the likelihood of vaccine uptake increased in G-7/F-7 compared to the control locality (AOR: 1.975, CI: 1.079, 3.617) but not anywhere else. Women were half as likely to express willingness to vaccinate but were not any different from men in terms of vaccine uptake. Increasing age, higher education and employment were important determinants of willingness and uptake of vaccination at the baseline. While these factors remained important at the endline as well, their significance decreased as seen by their lowered odds at the endline. Pashtuns became less likely and Urdu speakers more likely to receive vaccination at the endline.

A previous infection with COVID-19 for self was not a key determinant for willingness but a significant one for uptake of vaccination (AOR: 2.286, CI: 1.142, 4.576). Similarly, infection or vaccination of a family member and a high-risk perception were motivators for both willingness and uptake of vaccination. The effect of all these factors increased at endline. Having sought treatment for a recent illness was positively correlated with uptake (AOR: 1.471, CI: 1.078, 2.009).

Having received an SMS or call from the government was a major motivator that led to increased willingness (AOR: 2.414, CI: 1.420, 4.103) and uptake (AOR: 1.310, CI: 1.004, 1.708) in the endline. Advice from friends, family, medical professionals and religious leaders did not sway opinions about willingness or uptake of vaccination. Living near a CVC was correlated with higher willingness and closer distances were associated with higher uptake. The odds of vaccine uptake also increased if an NGO/CBO was working in the area in the pre-intervention period (AOR: 1.657, CI: 1.093, 2.514) (Table 5).

**Table 5.**
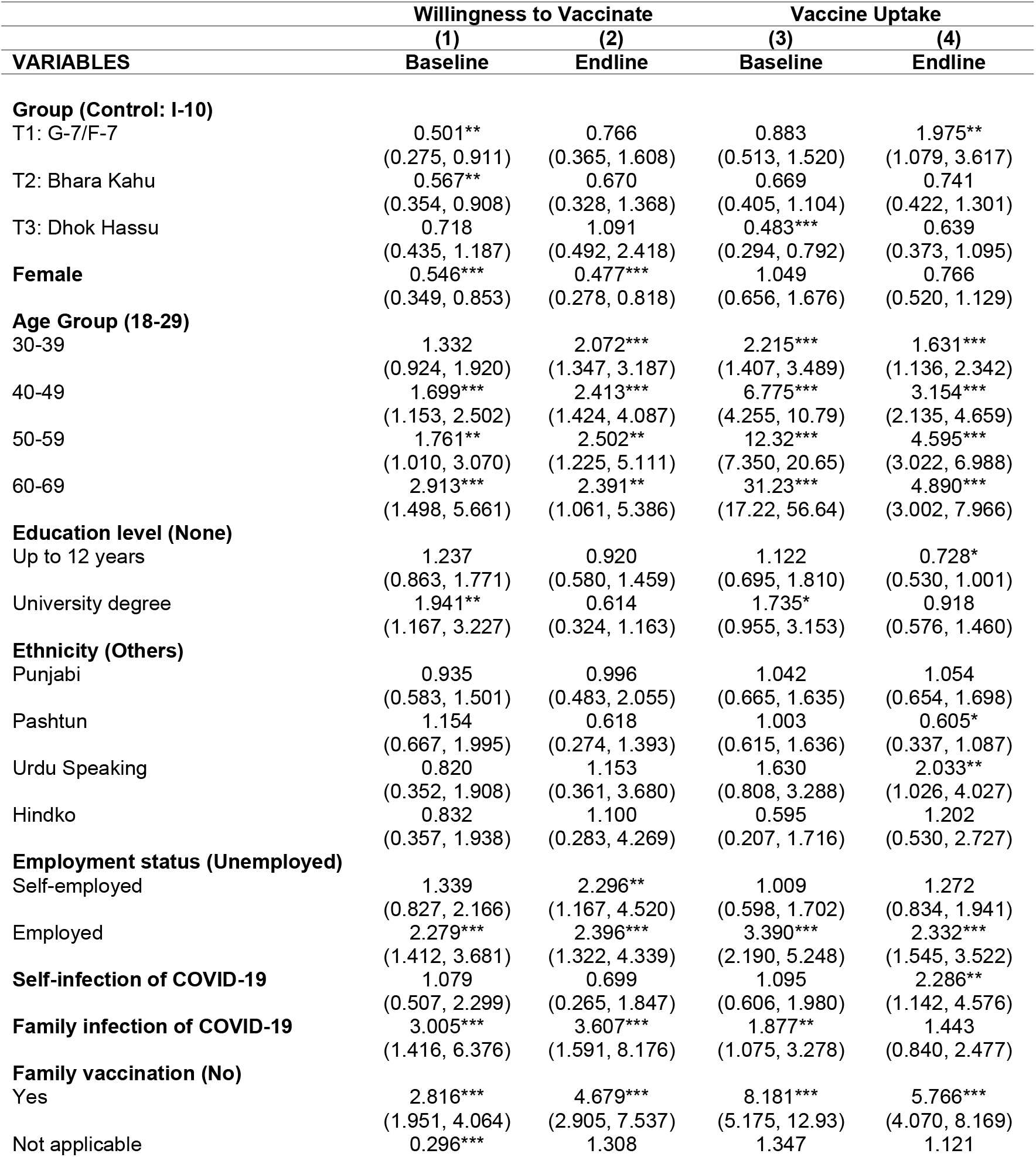
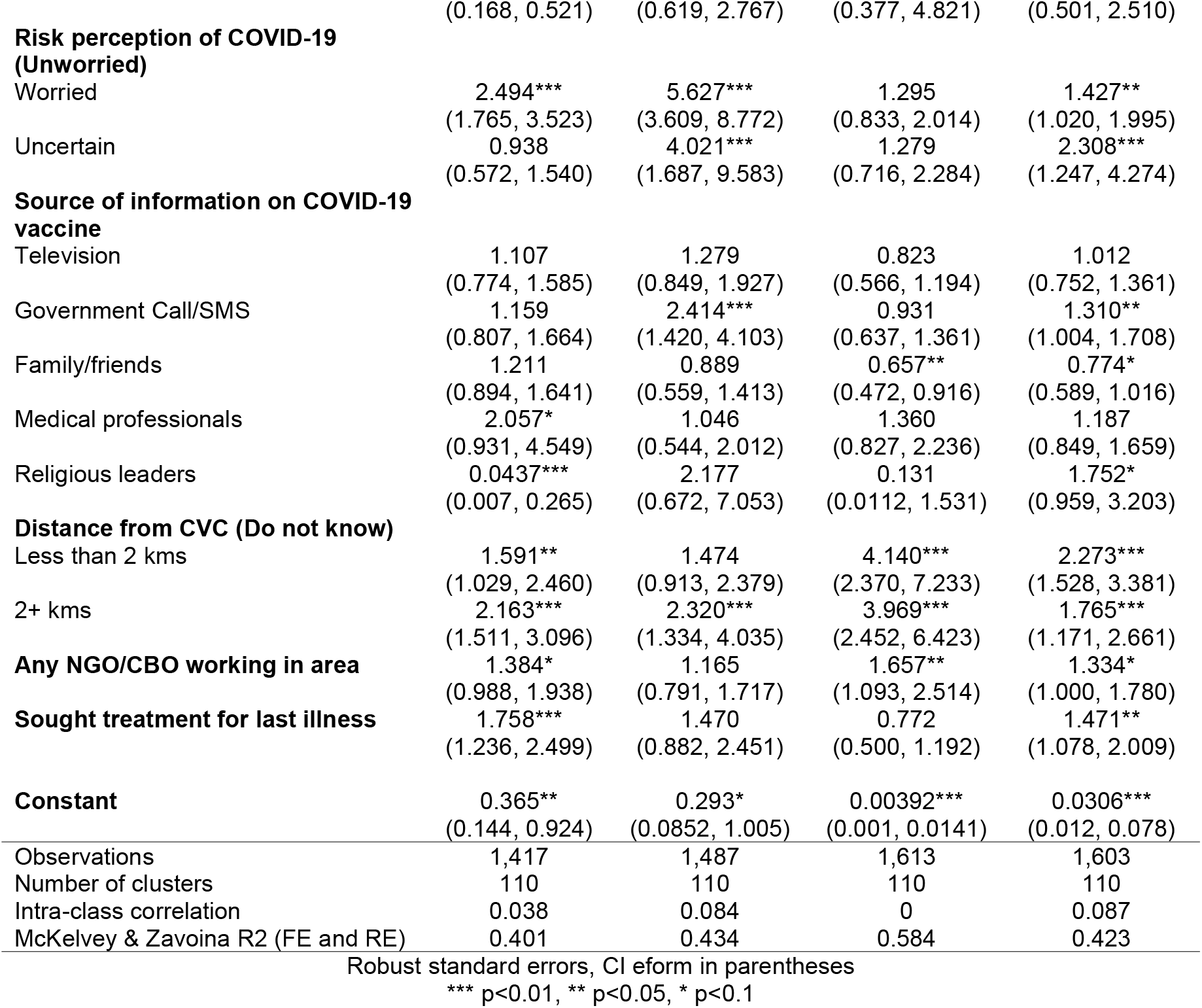
Odds ratios of factors influencing willingness to vaccinate and vaccine uptake.

## Discussion

We found that interventions that raise awareness and remove access barriers helped improve vaccine willingness by 7% and uptake by 17% in some low-income and underserved communities. However, there is a two-stage process. In the first, awareness increased and hesitancy decreased, following our awareness interventions. In the second stage, some of those that became convinced took up the vaccines. Uptake was dependent on access to vaccinations, which our interventions addressed only in part.

Our findings suggest that raising awareness of COVID-19 vaccination through more personalized means at community levels using printed material in local languages, engaging with community leaders, and building partnerships with local organizations can improve vaccine willingness by changing behavioral intentions of residents, which is in line with previous literature on the topic [11,29,32]. By contrast, merely informing the public through television, internet or local pamphlets etc., i.e., non-personalized means, may be less effective, as was seen in the control area. However, benefits from this approach may saturate beyond a certain point. For example, our interventions were successful in improving vaccine willingness in Bhara Kahu and Dhok Hassu by 7.6 and 6.6 percentage points compared to I-10, but to a lesser extent in G7/F7 where there had already been high willingness at baseline.

While willingness improved, it did not always translate into increased uptake. A major barrier towards achieving COVID-19 vaccination coverage in some settlements is the difficulty that residents have in accessing CVCs, which were often several kilometers away. While our set of interventions included some mobile vaccination centers (MVC), these were insufficient to fulfill the extent of demand for vaccination. Thus, vaccine uptake increased the most in G-7/F-7 by 17.1 percentage points compared to the control area, possibly because these areas are located in the center of Islamabad with easier access to multiple CVCs nearby – as compared to Bhara Kahu, a peri-urban slum outside Islamabad, and Dhok Hassu, an urban slum of Rawalpindi.

Table 5 tracks changes in willingness and uptake before and after the intervention. Odds of willingness to receive vaccination which was lower in all urban slums at baseline when compared to the control area, became indistinguishable at the endline. Similarly, the odds of willingness for university educated respondents, which were initially twice as much as those with lesser education, became insignificant. On the other hand, the odds of willingness rose for those that had experienced infection for self or family, if someone was already worried about infections or if a family member had received vaccination. It appears that the interventions may have helped mitigate the disadvantage of residence in an urban slum or from lower education and accentuated the willingness of those who had encountered infection or vaccines.

Similarly, odds of uptake of the vaccination rose by two-folds in G-7/F-7, which is located in city center and for the 18-39 years age groups compared to all older groups. On the other hand, there appeared to be little effect from education or employment and slight loss of advantage of those with a previous vaccination in the family. In short, in the endline period, there appears to be a homogenization effect in terms of who would take up vaccination, or a loss of disadvantage of the less educated, younger individuals, and residents of marginalized communities.

Our interventions also helped to increase vaccine willingness for people whose family members had a prior experience with COVID-19, but did not affect their uptake, which was more associated with a previous infection for self. Although our results are consistent with a prior study which uses a sample of Pakistan’s adult population to the extent that an incidence of COVID-19 among family members influences perception about COVID-19 vaccination [39], the effect is not strong enough to translate this willingness into action.

Vaccine uptake was higher among those with a previous infection with COVID-19, those with risk-aversion (were worried about getting infected) and those who sought treatment for their illness. Previous research has shown a positive association between health concerns and vaccine willingness [40–42], therefore our interventions may have nudged them to seek vaccination [43]. This implies that raising awareness, dispelling rumors, and communicating benefits of COVID-19 vaccines can change behavior of people who are more concerned about their health.

A prior study on routine immunizations in urban slums of Pakistan found that source of information also plays an important role in shaping trust and risk perceptions of vaccines [44]. Given the social norms and inaccurate information especially in low-income settlements, people might not get vaccinated due to social hesitancy regarding COVID-19 vaccination. We also found that government calls and SMS about COVID-19 vaccination in the endline were associated with both increased willingness and uptake. Given the high tele-density of cellphone users in Pakistan −85.3% penetration as of October 2021 [45], cellphone campaigns communicating COVID-19 and benefits of vaccination may be cost-effective.

### Limitations

One limitation is that a group (set) of interventions – community outreach through local leaders, local vaccination campaign and information -were implemented for all treatment groups. Therefore, we cannot isolate the effect of each intervention to analyze its relative effectiveness. Secondly, although MVCs were deployed in each treatment area and each reached around 150-250 vaccinations a day compared to 30-50 when they merely showed up without our intervention, such MVC visits were too few. Only 5% of the respondents out of total vaccinated reported that they had been vaccinated through MVCs, suggesting that CVCs were the predominant source of vaccinations and subject to the distance effect mentioned above. This may have limited the impact on vaccination uptake in distant communities (Bara Kahu and Dhok Hassu). Data for previous self and family infection of COVID-19 are self-reported and not actually verified through laboratory means which may have skewed our results to some extent. Finally, the baseline and endline were separated by 2 months, during which the national vaccination campaign had ramped up. Some of the homogenization of uptake may be explained by this rather an intervention effect, although the difference-in-differences analysis shows a significant change.

## Conclusion

We show that personalized information campaigns such as community mobilization and direct messaging are superior to general messaging in helping overcome COVID-19 vaccine hesitancy. However, increasing uptake of the vaccine requires an additional step of improving access to the vaccination services. Both information and access may be different for various communities and therefore a “one-size-fits-all” approach may need to be better localized. These findings may apply to other vaccinations and possibly to other health initiatives where the public may require motivation to uptake services such as diabetes or hypertension screening or testing. A key lesson is that low-income or marginalized communities would be better served if the services are brought to them locally.

## Data Availability

All relevant data are within the manuscript and its Supporting Information files.

## List of Legends

**S1 Fig. Pamphlet distributed for awareness campaign**

**S1 Table. Names, descriptions and coding of variables**

**S2 Table. Adjusted odds ratios for willingness to vaccinate and vaccine uptake**

**S1 Data. The dataset used for the study**

